# Presymptomatic plasma biomarkers in autosomal dominant Alzheimer’s disease: sequence and timing

**DOI:** 10.64898/2026.03.30.26349682

**Authors:** Christopher R S Belder, Amanda J Heslegrave, Owen Swann, Emily Abel, Millie Beament, Moneeb Nasir, Helen Rice, Philip S J Weston, Natalie S Ryan, Lyle J Palmer, Amy Brodtmann, Timothy Kleinig, Henrik Zetterberg, Nick C Fox

## Abstract

**Background:** Autosomal dominant Alzheimer’s disease (ADAD) serves as a model for presymptomatic biomarker discovery. Characterising the temporal profile of plasma biomarker levels in presymptomatic individuals may enhance understanding of disease pathogenesis, inform future clinical trials, and guide clinical interpretation.

**Methods:** We evaluated 124 proteins using a NUcleic acid-Linked Immuno-Sandwich Assay (NULISA) panel in 270 plasma samples from a longitudinal cohort study of ADAD, comprising 113 individuals (73 mutation carriers and 40 non-carriers). We determined the plasma proteomic changes that distinguished mutation carriers from non-carriers. We then used predicted age at symptom onset to determine the approximate timing of presymptomatic divergence in biomarker levels in carriers relative to non-carriers.

**Results:** Nine proteins (Aβ42, BACE1, GFAP, pTau181, pTau231, pTau217, MAPT, NfL, and AChE) robustly differed between carriers and non-carriers, cross-sectionally. Longitudinal analyses showed Aβ42 levels were elevated in carriers at least 26 years before expected symptom onset. Carriers diverged from non-carriers in phosphorylated tau markers at 21-24 years before expected symptoms, total-tau at 19 years, GFAP and BACE1 at 14 years, and NfL at 6 years. Differences in AChE were seen in symptomatic individuals, likely reflecting cholinesterase inhibitor use.

**Conclusion:** Multiple plasma proteins are elevated in presymptomatic and symptomatic autosomal dominant AD mutation carriers relative to non-carriers. Changes in eight biomarkers occur sequentially from 26 to 6 years prior to symptom onset. Combining biomarkers may help in staging presymptomatic AD and optimise clinical trial inclusion. Further work is needed to assess how these findings generalise to non-monogenic AD.

**What is already known on this topic:** The molecular pathology of Alzheimer’s disease develops many years before the onset of symptoms, and multiple plasma biomarkers of Alzheimer’s pathology have been identified. Understanding the timing of biomarker abnormality is important to guide trial design for the timing of interventions to prevent the onset of dementia.

**What this study adds:** Using an autosomal dominant Alzheimer’s disease cohort, we identify multiple plasma biomarkers that distinguish mutation carriers from non-carrier familial controls and characterise the timing of these changes relative to symptom onset. We demonstrate that biomarkers show change many years before symptom onset: markers of abnormal tau phosphorylation more than 20 years prior, followed by markers of reactive astrocytosis and synaptic dysfunction approximately 15 years prior, and neurodegenerative markers within 10 years of symptoms.

**How this study might affect research, practice or policy:** Plasma biomarkers could be used in pre-clinical autosomal dominant Alzheimer’s disease to chart disease trajectories and predict symptom onset, allowing targeted disease-modifying therapy implementation and optimised clinical trial design.

## Introduction

The molecular pathology of Alzheimer’s disease (AD) accumulates decades before symptom onset(1, 2). Multiple biomarkers associated with different aspects of AD pathology have been identified, including several well-validated blood-based biomarkers(1). However, the temporal sequence of presymptomatic changes in blood-based biomarkers is less well described.

Understanding the sequence, timing, and variability of biomarkers adds to the understanding of AD pathogenesis. It also has practical importance for presymptomatic trials that seek to delay or prevent clinical onset – a current critical priority in AD research(3).

Autosomal dominant Alzheimer disease (ADAD), caused by mutations in presenilin-1 (*PSEN1*), presenilin-2 (*PSEN2*) and amyloid precursor protein (*APP*), is a rare monogenic form of AD. Disease onset occurs at young ages, typically the fifth or sixth decade of life(4). The highly penetrant nature of ADAD mutations and relatively consistent variant-dependent age at onset (AAO)(4, 5) allows for the longitudinal study of individuals who carry pathogenic variants years or decades before the onset of symptoms. A person’s expected age at symptom onset can be calculated with reasonable precision based on the AAO of their affected parent or specific pathogenic variant(5). The typically young age also means there is less confounding from age-related co-pathology, allowing for higher confidence that observed changes relate to AD pathogenesis.

ADAD cohorts have been key to the discovery and validation of blood-based biomarkers of pre-symptomatic AD. Previous investigations have focused on single or a limited number of biomarkers(6-17), and have reported variable timings and sequences of change.

A major development in biomarker analytic technology has been the NUcleic acid-Linked Immuno-Sandwich Assay (NULISA)(18), which incorporates a ‘CNS panel’ of 124 analytes specifically of interest in neurodegenerative diseases, including high performance measurement of multiple phosphorylated tau species(19). This allows for the simultaneous quantification of many plasma biomarkers of importance in AD from small plasma volumes (∼30µl).

We leveraged the NULISA CNS panel to examine multiple plasma biomarkers in a cohort comprising both symptomatic and asymptomatic mutation carriers (MCs) of *PSEN1* and *APP* variants and compared their findings with well-matched non-carrier familial controls (NCs)(18). We aimed to examine the differential expression between MCs and NCs in plasma biomarker levels and evaluate their temporal changes relative to estimated age of symptom onset. By measuring multiple biomarkers in tandem from the same blood sample, using highly sensitive biomarker analytic technology and consistent statistical modelling approach, we sought to evaluate the relative timing of abnormality in these biomarker abnormalities to inform their use in AD molecular staging and prevention trials.

## Methods

### Study design and participants

We studied participants enrolled in a longitudinal cohort study of ADAD at the Dementia Research Centre, University College London. Cohort details have been described previously(20, 21).

Eligibility was either a clinical diagnosis of ADAD including confirmation of the presence of an ADAD causing mutation *or* being at risk of ADAD due to a genetically confirmed ADAD affected parent. ADAD mutation status was determined using Sanger sequencing; participants and study clinicians were blinded to genetic results. All participants with an appropriately stored research plasma sample were identified for potential inclusion in the current study. 120 potential participants were identified, with sample dates ranging from March 2018 to July 2024. Two participants with an *APP* duplication causing a predominant clinical manifestation of cerebral amyloid angiopathy (CAA) with intracerebral haemorrhage were excluded.

### Estimated years from onset (EYO)

Estimated years from onset was calculated i) in asymptomatic individuals by subtracting the AAO in the affected parent from the participant’s age at blood sampling and ii) in symptomatic individuals by subtracting the reported or observed AAO from age at blood sampling. Therefore, individuals with a negative EYO are younger than expected age at onset and those with a positive EYO are older. Individuals with inadequate information to determine EYO were excluded, as were NCs >20 years past EYO due to a lack of MCs with a comparable EYO (four individuals).

### Clinical assessments

Participants were followed annually (median number of visits=2, IQR=1-3; median interval between visits 1.06 years, IQR 0.98-1.46 years). At each visit a semi-structured health questionnaire and physical examination was conducted. Mini-Mental State Examination (MMSE)(22) and Clinical Dementia Rating (CDR) scale(23) were conducted according to standardised protocols at most visits (MMSE available for 231/270 visits, CDR for 214/270).

### Measurement of plasma proteins

Non-fasting plasma samples were collected from whole blood into EDTA-coated tubes at each visit. Samples were processed, centrifuged, aliquoted and frozen at -80_°_C according to standardised procedures. 49 individuals had 1 sample, 19 had 2, 17 had 3, 12 had 4, 12 had 5 and 4 had 6. 270 plasma samples were run in singlicate using the CNS 120 Panel (Supplementary Table 1) on the NULISA ARGO analyser platform (Alamar Biosciences)(18) at the UK Dementia Research Institute Biomarker Factory, London according to the manufacturer’s instructions. NULISA Protein Quantification (NPQ) values for each biomarker represent log_2_-transformed intensities that are normalised per plate using internal controls. One individual was excluded from subsequent analyses due to quality control failure during sample analysis.

**Table 1.** Participant characteristics.

|  | Non-carriers<br>n = 40 | Mutation carriers<br>n = 73 |  | p-value |
| --- | --- | --- | --- | --- |
|  |  | Asymptomatic<br>n = 43 | Symptomatic<br>n = 30 |  |
| Female sex, n (%) | 19 (47) | 27 (63) | 13 (43) | 0.199 |
| Age, years, mean (SD) | 36.6 (9.8) | 34.7 (8.0) | 50.4 (8.9) | <0.001<sup>a</sup> |
| Mutation |  |  |  | 0.967 |
| *PSEN1*, n (%) | 27 (67) | 30 (70) | 21 (70) |  |
| *APP*, n (%) | 13 (33) | 13 (30) | 9 (30) |  |
| EYO, years, mean (SD) | –11.0 (10.9) | –13.8 (8.4) | 4.9 (3.1) | <0.001<sup>a</sup> |
| *APOE*-ε4 carrier, n (%) | 13 (32) | 12 (28) | 9 (30) | 0.901 |
| MMSE, Median (IQR) | 30 | 30 | 23 (17-24) | <0.001<sup>a</sup> |
| CDR Global, Median (IQR) | 0 | 0 | 0.5 (0.5-1) | <0.001<sup>a</sup> |
| CDR-SB, Median (IQR) | 0 | 0 | 4 (1-7) | <0.001<sup>a</sup> |
| Number of visits, median (range) | 2 (1-6) | 2 (1-6) | 1 (1-4) |  |

<sup>a</sup> Significant difference between non-carriers vs. symptomatic mutation carriers and asymptomatic vs. symptomatic mutation carriers. *PSEN1* – Presenilin 1. *APP* – Amyloid Precursor Protein. *APOE* – Apolipoprotein E. EYO – Estimated Years from Onset. MMSE – Mini Mental State Examination. CDR – Clinical Dementia Rating. CDR-SB – CDR Sum of Boxes.

### Statistical analysis

#### Cross-sectional analyses

Visit data and blood samples from the earliest visit per individual were included for the cross-sectional analyses. Participant characteristics are presented as mean and standard deviation (SD) or median and interquartile range (IQR) for continuous variables and n (%) for categorical variables.

p-values were determined using Chi-squared tests for categorical variables, and analysis of variance (ANOVA) or Kruskal-Wallis tests for continuous variables, with post-hoc Tukey’s and Dunn tests as appropriate.

For all analytes, linear regression models were used to compare sex-adjusted protein levels between the MC and NC groups. The inherently log_2_-transformed NPQ model outputs were back-transformed to report the mean fold-change (FC) in protein level between groups. Sensitivity analyses were conducted by excluding extreme outliers (±3.0 × interquartile range (IQR) for each protein) prior to model fit and adjusting for age by including this as a covariate.

To explore differences in protein expression across different temporal stages, the cohort was partitioned into three EYO based groups: EYO ≥0, and those with EYO <0 were divided in two groups separated by the median EYO – the first group comprising EYO −14.7 to 0 (closer to expected symptom onset) and the second individuals with EYO <−14.7 (further from expected onset). Mean protein levels in the MC group were compared to the NC group in each of these 3 strata, adjusting for age and sex.

To investigate mutation-specific associations, mean protein levels were compared between carriers of *PSEN1* and *APP*, adjusting for age, EYO and sex. Separate comparisons of *PSEN1* MCs and *APP* MCs groups with pooled NCs were also performed, adjusting for age and sex.

Estimated p-values were adjusted for multiple testing using the false discovery rate (FDR) method(24).

#### Longitudinal analyses

Using available samples from all study visits for each participant, we sought to evaluate a pre-specified subset of established plasma proteins and their temporal change relative to EYO, as well as any additional markers of interest identified in the cross-sectional analyses. The pre-specified subset comprised those highlighted in the 2024 revised criteria for diagnosis and staging of AD(1): tau phosphorylated at threonine 181 (pTau181), threonine 217 (pTau217), and threonine 231 (pTau231); glial fibrillary acidic protein (GFAP) and neurofilament light (NfL). The association of mutation status with biomarker level relative to EYO was assessed using linear mixed effects (LME) models, incorporating restricted cubic splines to account for potential non-linear relationships with knots at the 10^th^, 50^th^ and 90^th^ quantiles(25). EYO, mutation status, a mutation status:EYO interaction term, age, and sex were modelled as fixed effects. Random effects for ‘individual’ and ‘family’ were incorporated to account for hierarchical clustering. To estimate the timing of divergence in biomarker level in MCs relative to NCs, the LMEs were used to compute marginal means for MCs and NCs at each integer EYO, the difference and 95% CIs between these were calculated, and the EYO at which the 95% CI did not include zero was taken as the timepoint of divergence(9, 14, 21). Extreme outliers for each biomarker (±3.0×IQR), if present, were excluded prior to model fit. To evaluate the impact of the chosen significance threshold on the timepoint of divergence, estimates using 90% and 99% CIs were also generated. Statistical analyses were performed using Rv4.4.2. The STROBE reporting guideline was used to draft this manuscript(26).

## Results

Baseline demographic and clinical characteristics are presented in Table 1. A total of 113 individuals with 270 plasma samples were included in the final analysis, comprising 73 MCs, 30 were symptomatic (57 samples) and 43 asymptomatic (108 samples) at baseline, and 40 NC familial controls (105 samples). Two MCs who were initially asymptomatic became symptomatic between visits. A total of 38 distinct ADAD causing variants were represented in the families in our cohort, comprising 7 *APP* and 31 *PSEN1* variants (Supplementary Table 2).

### Cross-sectional analyses

Comparing MCs vs NCs, nine proteins were identified as differentially expressed after FDR correction: pTau217 (Fold-change 2.63, FDR<0.001), pTau231 (FC 2.54, FDR<0.001), and pTau181 (FC 2.17, FDR<0.001); GFAP (FC 2.03, FDR<0.001), total tau (MAPT) (FC 1.69, FDR<0.001), amyloid-beta 42 (Aβ42) (FC 1.70, FDR<0.001), NfL (FC 1.60, FDR<0.001), beta-secretase (BACE1) (FC 1.17, FDR=0.004), and acetylcholinesterase (AChE) (FC 1.30, FDR=0.016) (Figure 1). The identified proteins were robust to sensitivity analyses, excepting AChE, which was no longer significant after FDR correction when age was included as a covariate.

**Fig. 1.**
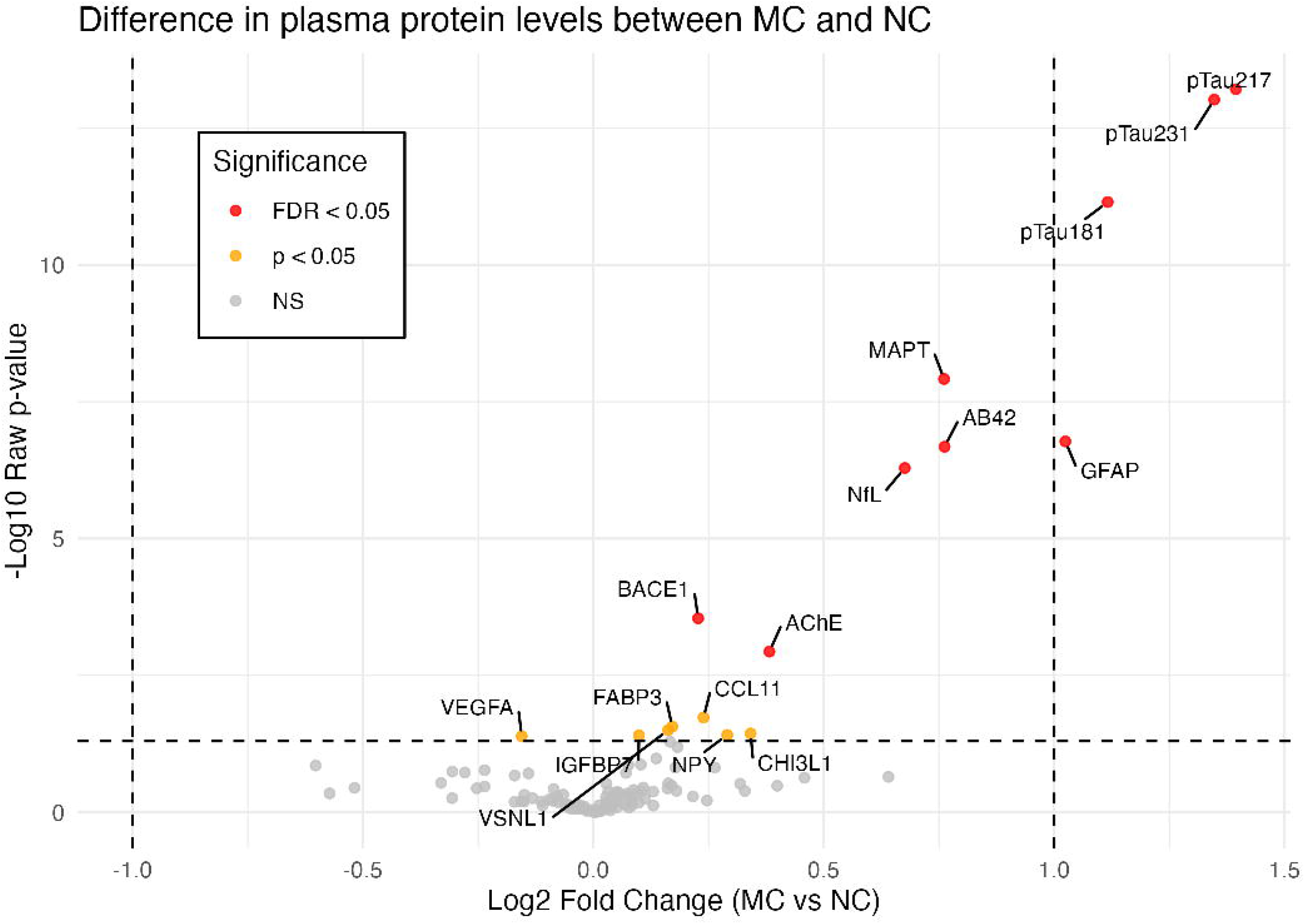

Distinct proteins were identified as differentially expressed across EYO groups in the exploratory analyses (Figure 2A-C). In individuals with EYO≥0, (past expected symptom onset or symptomatic, n=40), pTau217, pTau231, pTau181, GFAP, MAPT and NfL remained the most elevated in MCs vs NCs, consistent with the findings in the complete cohort. In the late-presymptomatic stage (EYO −14.7-0, n=37), phosphorylated tau species, GFAP and Aβ42 were identified as the most elevated in MCs vs NCs, with additional increased VSNL1 (FC 1.42, FDR=0.006) and TEK (FC 1.22, FDR=0.016) and reduced PDLIM5 (FC 0.26, FDR=0.023). In the early presymptomatic stage (EYO < −14.7, n=36), no analytes were significantly different after FDR correction. Among the nominally significant proteins, Aβ42 had the greatest increase (FC 1.36, p=0.008), followed by pTau231 (FC 1.29, p=0.013).

**Fig. 2.**
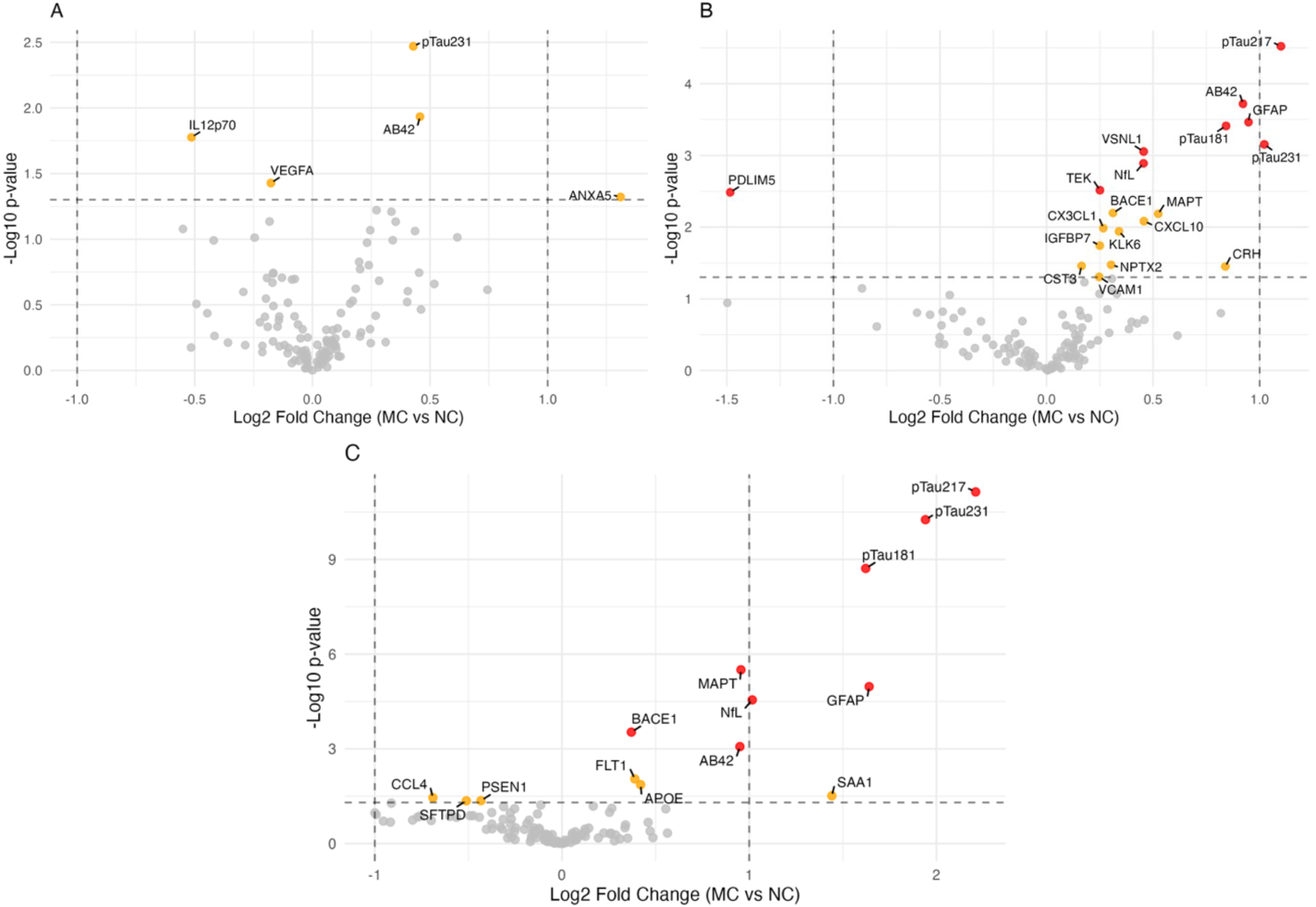

In the exploratory comparison between *PSEN1* and *APP* MCs (Figure 3A), Aβ38 demonstrated the greatest difference between groups, being increased in *APP*-MCs relative to *PSEN1*-MCs (FC 2.25, FDR <0.001), while Aβ40 (FC 1.37, FDR=0.013), was increased in *PSEN1* relative to *APP* MCs.

**Fig. 3.**
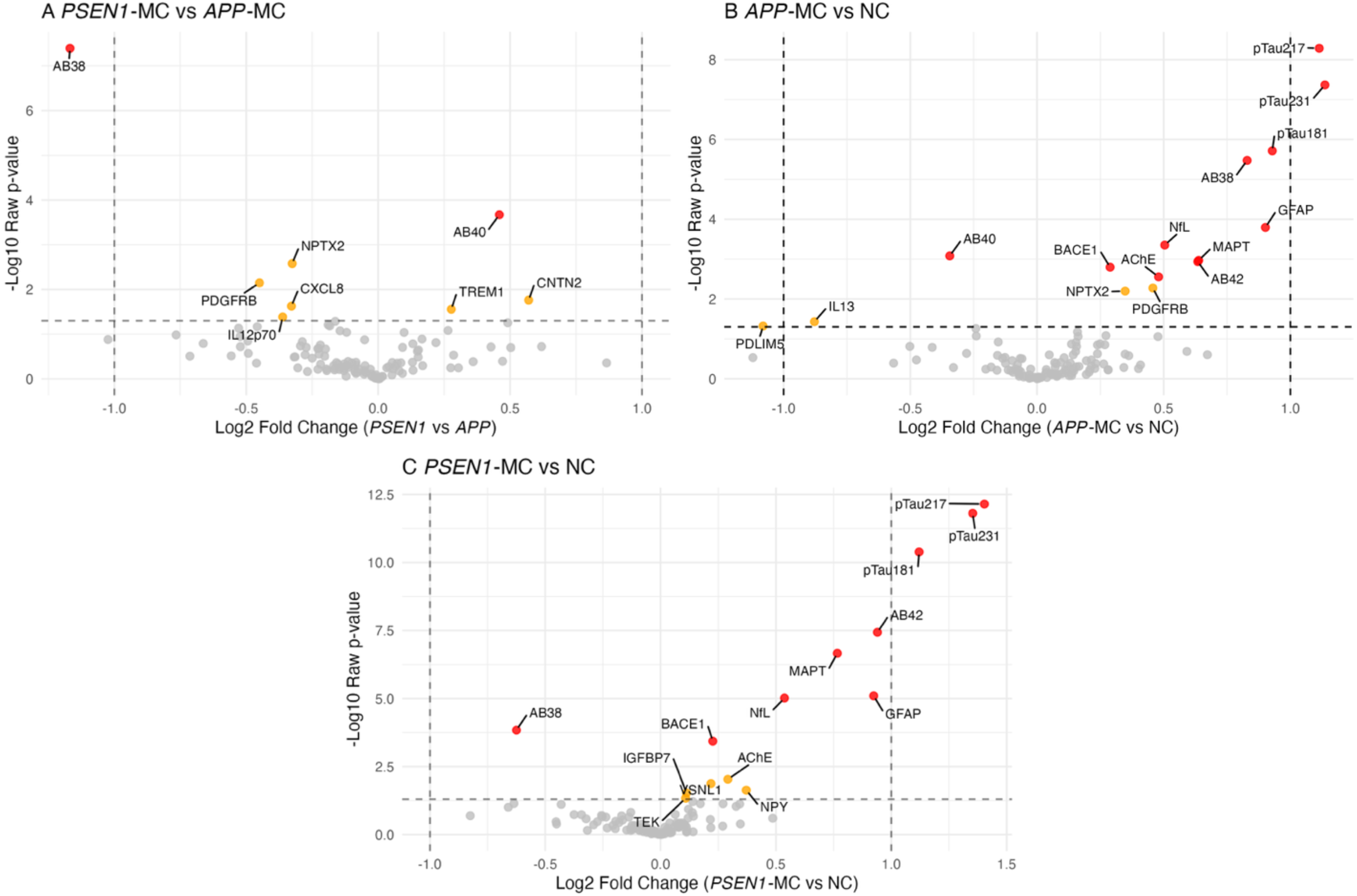

Comparing the two MC groups of *APP* and *PSEN1* carriers separately with pooled NCs identified the same proteins as in the complete MC vs NC analysis (excepting AChE for *PSEN1*-MCs vs NCs as nominally significant only), as well as additional findings (Figure 3B,C): In *APP* MCs vs NCs (n=63), Aβ40 was reduced (FC 0.79, FDR=0.001) and Aβ38 was elevated (FC 1.78, FDR<0.001), whereas in *PSEN1* MCs vs NCs (n=93), Aβ38 was lower in MCs than NCs (FC 0.65, FDR=0.002). Nominally significant findings are included in figures for completeness.

### Longitudinal analyses

In the longitudinal analyses, Aβ42 demonstrated the earliest changes relative to expected symptom onset, being higher in MCs than NCs from EYO ≈ −26 years (α=90%: −27 years, α=99%: −24 years). The difference between MCs and NCs in Aβ42 levels was observable from the earliest EYO, although was not statistically discernible at these early timepoints due to a limited number of observations (Figure 4A). Phosphorylated tau markers and total-tau demonstrated the next earliest timing of divergence between the MC and NC groups: pTau231 at EYO ≈ −24 years (α=90%: −25, α=99%: −21), pTau181 at EYO ≈ −23 years (α=90%: −25, α=99%: −20), pTau217 at EYO ≈ −21 years (α=90%: −23, α=99%: −19), and total-tau (MAPT) at EYO ≈ −19 years (α=90%: −21, α=99%: −16) (Figure 4B-E). Later divergence was observed in GFAP at EYO ≈ −14 years (α=90%: −15, α=99%: −12), BACE1 at EYO ≈ −14 years (α=90%: −15, α=99%: −9), and then NfL at EYO ≈ −6 years (α=90%: −7, α=99%: −5) (Figure 4F-H).

**Fig. 4.**
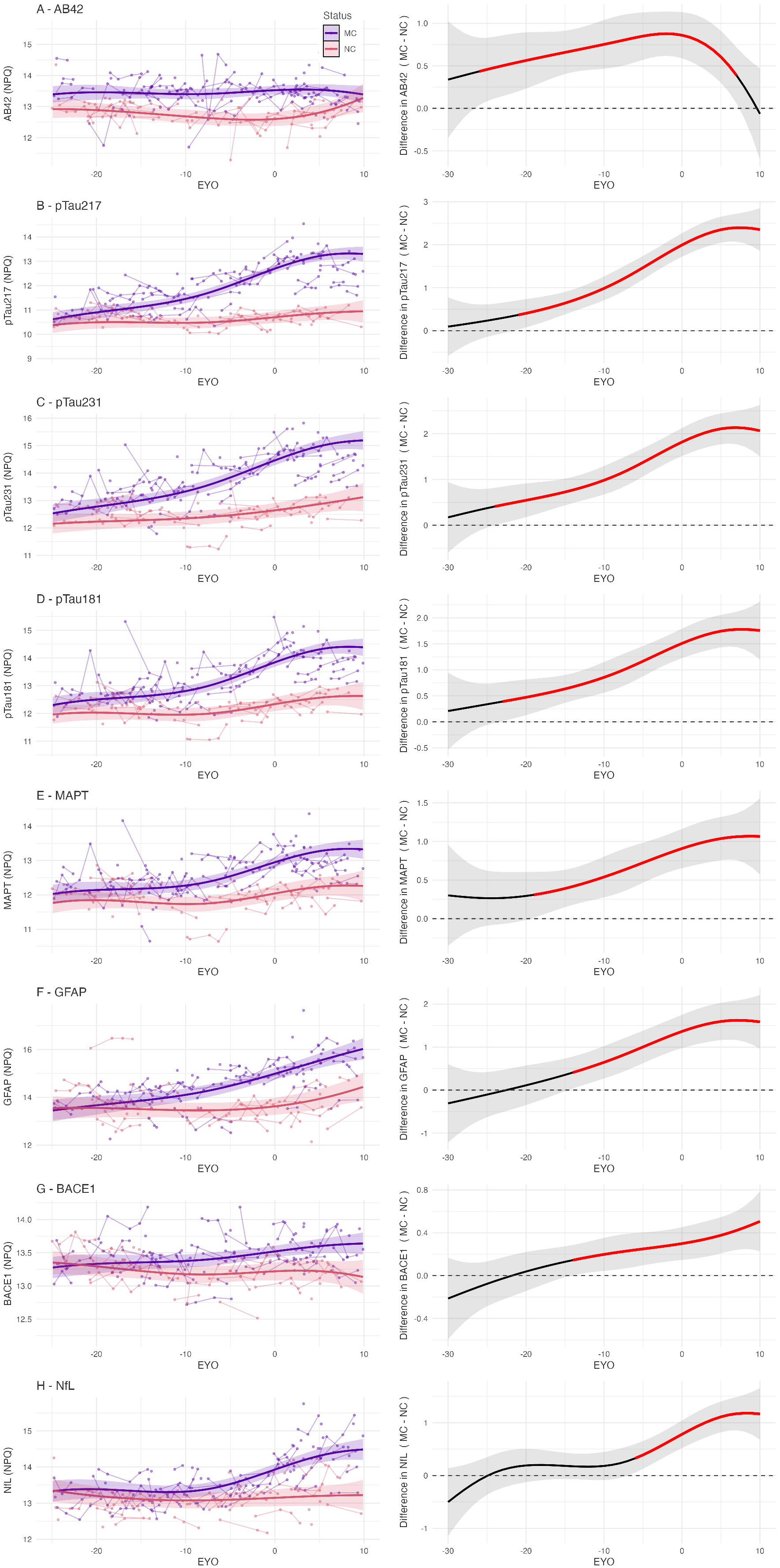

**Fig. 5.**
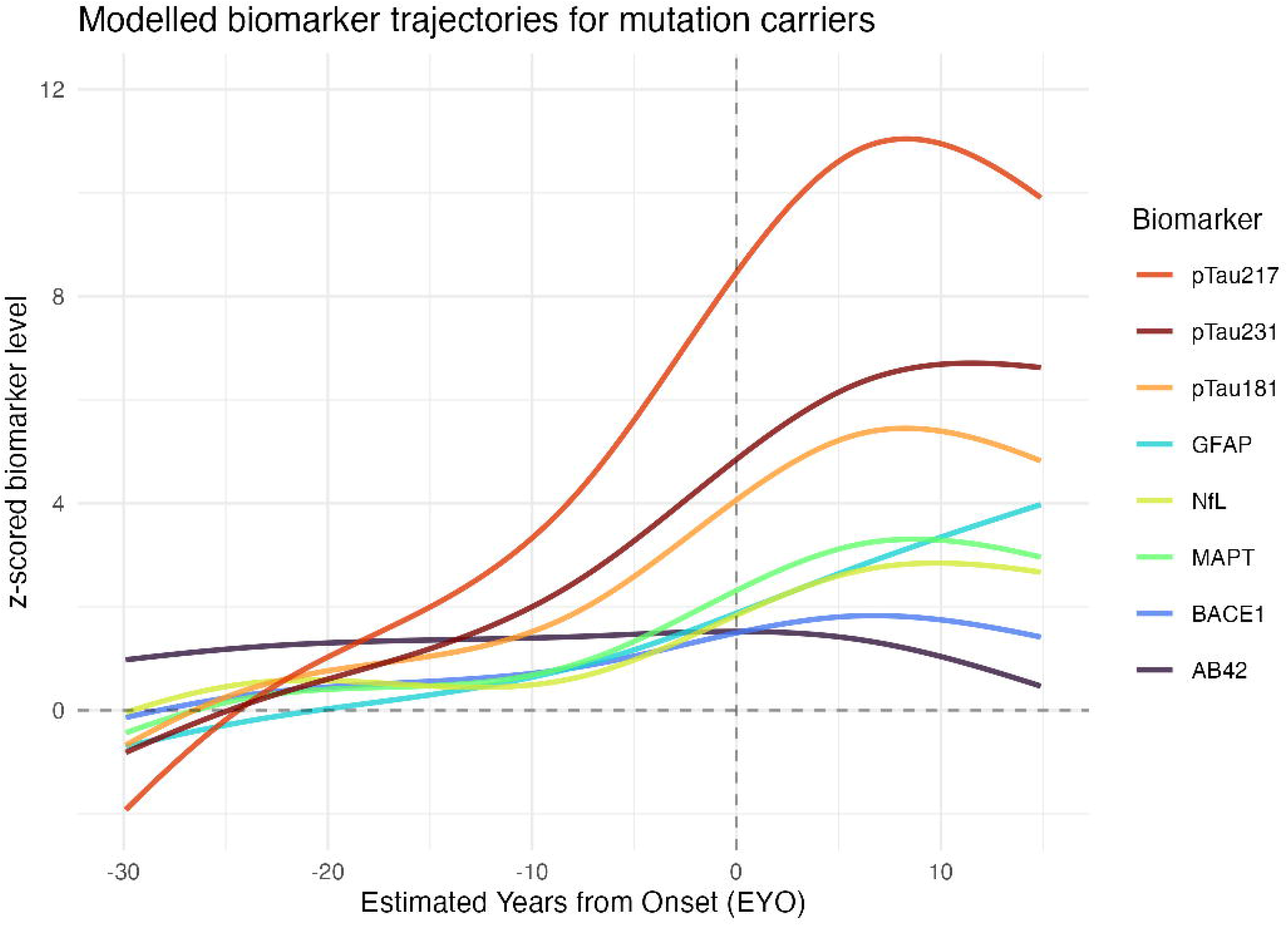

AChE appeared to diverge between MCs and NCs around symptom onset (EYO ≈ 0) (supplementary figure 1A). Additional analyses were performed examining participants for whom cholinesterase inhibitor (ChI) status was known (n = 94). When the longitudinal analysis was repeated restricted to the cohort known to not be prescribed ChI, there was no difference between MC and NC at any timepoint (supplementary figure 1B). In this subgroup, the baseline AChE level was higher in symptomatic individuals prescribed ChI compared with those not prescribed ChI, as well as asymptomatic mutation carriers and non-carriers (supplementary figure 1C).

## Discussion

In this observational study of plasma biomarkers in a longitudinal cohort of ADAD families, we report cross-sectional and longitudinal changes in a large panel of plasma biomarkers. We identified multiple plasma biomarkers that are elevated in ADAD MCs compared to NC familial controls and demonstrated their relative timing of divergence compared with estimated symptom onset. Both cohort-wide and gene-specific abnormalities were detected. The sequential biomarker abnormalities observed support distinct pathophysiological phases – initial abnormal production or metabolism of amyloid-beta, followed by markers of accumulating downstream cerebral pathology.

Elevated Aβ42 was identified as the earliest marker that discriminates between MCs and NCs, with statistically discernible difference 26 years before estimated symptom onset, and a nominal difference from the earliest observed EYO within our cohort (Figure 4A). Previous studies have supported this as a very early change, including elevated plasma Aβ42 and Aβ42/40 ratio even in childhood in *PSEN1* E280A carriers using a multiplexed fluorometric bead-based immunoassay(15, 27). These findings are consistent with ADAD pathogenic variants causing a constitutive alteration in APP processing, with an increased production of aggregation prone forms such as Aβ42, which is detectable in plasma. This is concordant with a prior study in our cohort, which found elevated plasma Aβ42/40 ratios in MCs using immunoprecipitation liquid chromatography-tandem mass spectrometry (IP-LC-MS/MS)(6).

There was a significant difference in Aβ38 and Aβ40 levels between individuals carrying *APP* and *PSEN1* variants. Aβ38 was elevated in *APP*-MCs (with a greater relative increase than Aβ42) and reduced in *PSEN1*-MCs relative to NCS while AB40 was reduced in *APP* MCs compared with *PSEN1*-MCs and NCs. Elevated Aβ38 (alongside Aβ40 and Aβ42) has been reported in *APP* Swedish (*APP* KM670/671NL) variant carriers, and low Aβ38 in *PSEN1* H163Y carriers (15).

Elevated Aβ42/38 has been observed in *PSEN1* MC vs *APP* MC and NC, and elevated Aβ38/40 in *APP* MC vs *PSEN1* MC and NC(6). These early changes in ADAD, likely representing mutation-specific effects on Aβ peptide production, are distinct from those seen in sporadic AD, where a reduced Aβ42/40 ratio is seen cerebrospinal fluid (CSF) (and to a lesser extent in plasma) with cerebral amyloid accumulation(28).

Phosphorylated tau species demonstrated both early temporal change, approximately 20 years before symptom onset, and the greatest relative increase in plasma levels between MCs and NCs. Our estimated timing of abnormality relative to EYO for pTau217 is consistent with findings in a large cohort of *PSEN1* E280A families at approximately 20 years before onset(16). The timing of difference in pTau181 is earlier than previous published analyses, which suggested divergence could be detected ranging from 6-16 years before onset of symptoms(14, 21). These disparate results may be accounted for by our larger sample size and the potentially more sensitive measurement technique. Acknowledging the imprecision associated with our relatively small sample size, the point estimates suggest that pTau231 and pTau181 become abnormal at an earlier EYO than pTau217 (at approximately −24 and −23 vs −21 years respectively). This accords with previous analyses in sporadic AD which suggested plasma pTau231 may be a marker of early change(29, 30), and a recent investigation in an ADAD cohort of PSEN1 E280A, which found earlier change in pTau231 than pTau217 when measured using Single molecule array (Simoa) immunoassay(31).

Elevations in plasma GFAP were observed at an intermediate time point: after increases in phosphorylated tau but before neurodegeneration (as indicated by NfL). The estimated timing of approximately 14 years before onset is consistent with previous reports examining blood GFAP in isolation in ADAD, which reported divergence between MCs and NCs at 10(11) and 16 years(10) before onset. Our sequence differs from the findings of Johansson et al.(14), who measured GFAP using Simoa, and found change at a similar 10 years before onset, but found this to occur earlier than both pTau181 and NfL (at 6 and 2 years before onset respectively). Johansson et al. included a smaller number of individuals (33 MCs, 42 NCs) and only a limited number of variants, with the Swedish and Arctic *APP* variants comprising a large proportion of individuals. These variants lead to particularly severe CAA as well as amyloid plaques(32), which may also contribute to elevated plasma GFAP(33). Modelling in sporadic AD populations relative to amyloid- and tau-PET positivity has also proposed plasma GFAP as an early change, however with high inter-individual variability(34).

Elevations in plasma NfL were seen in closest proximity to symptom onset, consistent with its status as a marker of neuronal injury. In previous papers examining serum and plasma NfL in ADAD cohorts, there have however been very different reports of the timing of abnormality, from 2 years before predicted onset(14), ranging through 7(8) and 19(9) to as early as 22 years before onset(17). This may perhaps be accounted for by the non-linear relationship between NfL and EYO we observed (Figure 4H). In a large cohort, some group separation might be observed very early, contingent on the modelling approach and threshold of significant difference chosen between groups, however a significant group separation is not seen until closer to symptom onset(17).

Plasma total-tau elevation has not, to our knowledge, been reported in ADAD previously. Studies using Simoa to measure plasma t-tau did not detect a difference between MCs and NCs(12, 14). This could be explained by greater power due to the larger sample size in our analysis, or the epitope targeted by the NULISA assay may differ from Simoa. Brain-derived tau (BD-tau) is likely to be still more informative(35), however this was not available on NULISA at the time of this analysis.

Our finding of elevated plasma BACE1 in ADAD MCs has not previously been reported, but has been observed in sporadic AD and MCI(36). The intermediate timing of abnormality seen here, approximately 14 years before onset, suggests that rather than being anchored to the early changes in amyloid processivity due to ADAD pathogenic variants, elevated BACE1 in this context may act as a marker of synaptic injury, as it is mostly expressed in presynaptic terminals(37).

Elevated AChE in symptomatic mutation carriers relative to controls was most likely due to ChI treatment. This highlights an important interpretive and analytic consideration when performing proteomic studies – that some findings may be a consequence of treatment rather than representing pathophysiologic changes.

In addition to the changes in amyloid processing, our exploratory analyses identified other plasma proteins which nominally differed between *PSEN1* and *APP* MCs. *PSEN1* mutations alter gamma secretase function and so may lead to altered of processing of other gamma secretase substrates, which in turn may contribute to the atypical phenotypes seen with *PSEN1* variant carriers such as spastic paraparesis(38). These findings warrant investigation in larger cohorts.

Our results have several important clinical and research implications. We reinforce phosphorylated tau markers as being the highest performing markers in distinguishing AD MCs from controls and highlight that elevation can be seen many (>20) years before symptom onset. Alterations in plasma amyloid-beta profiles, due to changes in APP processivity in ADAD, highlight the different considerations in biomarker interpretation between ADAD and sporadic AD. Importantly, elevated plasma Aβ42 could lead to a false negative plasma pTau217/Aβ42 ratio(39) in an individual being investigated for potential young onset AD if they harbour an ADAD pathogenic variant. By simultaneously examining multiple blood biomarkers within the same cohort, we provide further weight to the temporal sequence of proposed core and additional biomarkers of AD, with initial changes in pTau, followed by synaptic and glial markers (GFAP, BACE1), and late changes in markers of neuronal injury (typified by NfL).

## Limitations

Limitations of the current study include potential unmeasured confounders, including pre-analytic factors such as non-fasting sample collection and patient factors such as body-mass index and kidney or liver function, known to impact some plasma protein levels. Given the rarity of ADAD, our sample size was limited. However, our estimates of timing of changes in our longitudinal analyses are similar to previous analyses in larger cohorts reporting select individual biomarkers, supporting the robustness of our results. A further limitation is the absence of CSF or PET markers of amyloid or tau pathology. Validation in additional ADAD cohorts and in cohorts of sporadic AD and ageing will be important.

## Conclusions

Our current work expands our understanding of presymptomatic biomarker changes in AD and suggests the potential of using biomarker combinations measured from a single sample to improve diagnostic and predictive performance. In trials targeting presymptomatic AD pathology, using multiple biomarkers may help stage individuals and aid in estimating future risk of cognitive impairment, proximity to symptom onset and the tailoring of pharmacologic and non-pharmacologic interventions.

## Data Availability

To protect confidentiality of participants, data will not be made publicly available as complete anonymisation cannot be guaranteed due to the nature of the cohort. Anonymised data will be shared only if requested by a qualified academic investigator for replication of procedures and results presented and if data transfer is in agreement with legislation on general data protection regulation and supported via a material transfer or data processing agreement as appropriate.

## Ethics Statements

### Patient consent for publication

Not applicable

### Ethics approval

This study involves human participants and was approved by local research ethics committees (Queen Square Ethics Committee 11/LO/0753; University of Adelaide H-2025-042). Written informed consent was obtained from all participants, or from a surrogate if cognitive impairment precluded informed consent.

## Acknowledgements

We thank the participants and their families for their generous support of this study.

