## Supplementary Figures and Tables for "Presymptomatic plasma biomarkers in autosomal dominant Alzheimer’s disease: sequence and timing"

**Supplementary Figures and Tables to: Presymptomatic plasma biomarkers in autosomal dominant Alzheimer’s disease: sequence and timing**

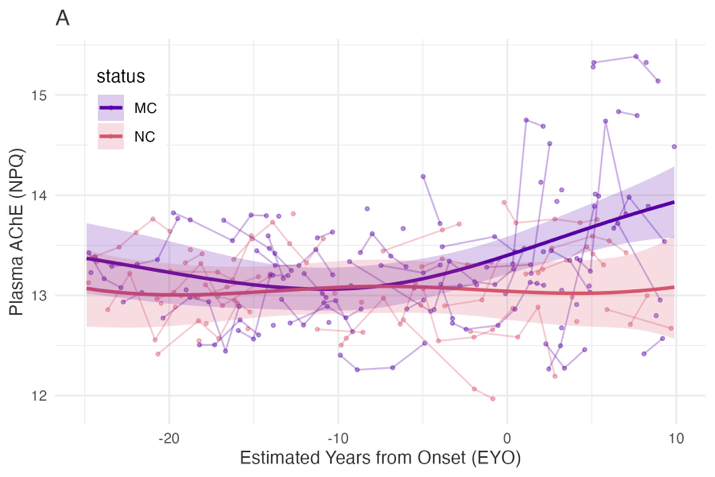

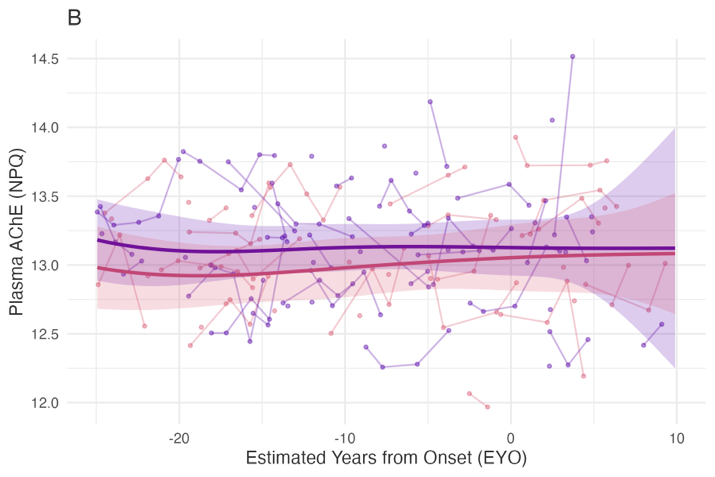

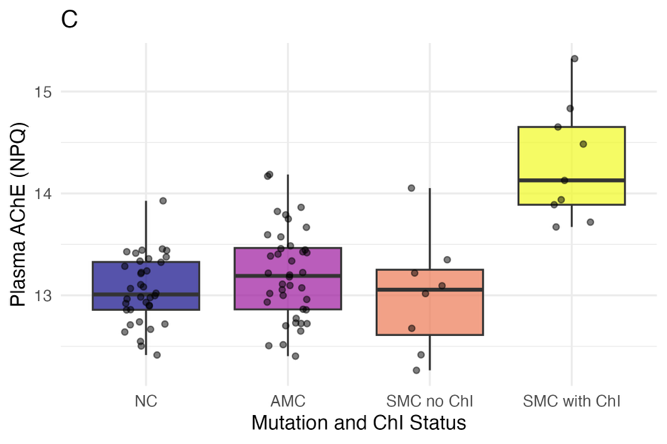

**Supplementary Figure 1 – Plasma acetylcholinesterase (AChE) levels by estimated years from onset and clinical status.** A and B - Coloured lines represent model-based predictions from linear mixed models, and shaded ribbons represent 95% CIs, stratified by mutation status. Observed data-points are superimposed with lines connecting repeated measures per individual. X axes are truncated and jitter has been applied to prevent inadvertent identification of individuals. A – full longitudinal cohort. B – cohort not taking cholinesterase inhibitors (ChI). C – box plots comparing cross-sectional plasma AChE levels stratified by mutation and ChI status with datapoints superimposed with jitter. MC: mutation carrier. NC: non-carrier. AMC: asymptomatic mutation carrier. SMC: symptomatic mutation carrier.

**Supplementary Table 1 – Proteins analysed by the NULISA CNS 120 Panel in this analysis**

| **Symbol** | **Full Protein Name** |
| --- | --- |
| AChE | Acetylcholinesterase |
| AGRN | Agrin |
| ANXA5 | Annexin A5 |
| APOE | Apolipoprotein E |
| APOE4 | Apolipoprotein E isoform 4 |
| ARSA | Arylsulfatase A |
| Aβ38 | Amyloid-beta precursor protein (Aβ38) |
| Aβ40 | Amyloid-beta precursor protein (Aβ40) |
| Aβ42 | Amyloid-beta precursor protein (Aβ42) |
| BACE1 | Beta-secretase 1 |
| BASP1 | Brain acid-soluble protein 1 |
| BDNF | Brain-derived neurotrophic factor |
| CALB2 | Calretinin (Calbindin 2) |
| CCL11 | Eotaxin (C-C motif chemokine ligand 11) |
| CCL13 | C-C motif chemokine ligand 13 |
| CCL17 | C-C motif chemokine ligand 17 |
| CCL2 | C-C motif chemokine ligand 2 |
| CCL22 | C-C motif chemokine ligand 22 |
| CCL26 | C-C motif chemokine ligand 26 |
| CCL3 | C-C motif chemokine ligand 3 |
| CCL4 | C-C motif chemokine ligand 4 |
| CD40LG | CD40 ligand |
| CD63 | Tetraspanin-30 / CD63 |
| CHI3L1 | Chitinase-3-like protein 1 |
| CHIT1 | Chitotriosidase-1 |
| CNTN2 | Contactin-2 |
| CRH | Corticotropin-releasing hormone |
| CRP | C-reactive protein |
| CSF2 | Granulocyte-macrophage colony-stimulating factor (GM-CSF) |
| CST3 | Cystatin C |
| CX3CL1 | Fractalkine (C-X3-C motif chemokine ligand 1) |
| CXCL1 | Growth-regulated alpha protein / C-X-C motif chemokine ligand 1 |
| CXCL10 | C-X-C motif chemokine ligand 10 (IP-10) |
| CXCL8 | Interleukin-8 (C-X-C motif chemokine ligand 8) |
| ENO2 | Gamma-enolase / Enolase 2 |
| FABP3 | Fatty acid-binding protein, heart / FABP3 |
| FCN2 | Ficolin-2 |
| FGF2 | Fibroblast growth factor 2 |
| FLT1 | Vascular endothelial growth factor receptor-1 (Fms-related tyrosine kinase 1) |
| FOLR1 | Folate receptor alpha |
| GDF15 | Growth/differentiation factor 15 |
| GDI1 | RuvB-like GTP-dissociation inhibitor 1 |
| GDNF | Glial cell line-derived neurotrophic factor |
| GFAP | Glial fibrillary acidic protein |
| GOT1 | Aspartate aminotransferase, cytoplasmic / Glutamic-oxaloacetic transaminase 1 |
| HBA1 | Hemoglobin subunit alpha |
| HTT | Huntingtin |
| ICAM1 | Intercellular adhesion molecule 1 |
| IFNG | Interferon gamma |
| IGF1R | Insulin-like growth factor 1 receptor |
| IGFBP7 | Insulin-like growth factor-binding protein 7 |
| IL10 | Interleukin-10 |
| IL12p70 | Interleukin-12 (p70 heterodimer) |
| IL13 | Interleukin-13 |
| IL15 | Interleukin-15 |
| IL16 | Pro-interleukin-16 |
| IL17A | Interleukin-17A |
| IL18 | Interleukin-18 |
| IL1B | Interleukin-1 beta |
| IL2 | Interleukin-2 |
| IL33 | Interleukin-33 |
| IL4 | Interleukin-4 |
| IL5 | Interleukin-5 |
| IL6 | Interleukin-6 |
| IL6R | Interleukin-6 receptor subunit alpha |
| IL7 | Interleukin-7 |
| IL9 | Interleukin-9 |
| KDR | Vascular endothelial growth factor receptor-2 (VEGFR-2) |
| KLK6 | Kallikrein-related peptidase 6 |
| MAPT | Microtubule-associated protein tau |
| MDH1 | Malate dehydrogenase 1, cytoplasmic |
| MME | Membrane metalloendopeptidase / Neprilysin |
| MSLN | Mesothelin |
| NEFH | Neurofilament heavy polypeptide |
| NEFL | Neurofilament light polypeptide |
| NGF | Beta-nerve growth factor / Nerve growth factor |
| NPTX1 | Neuronal pentraxin-1 |
| NPTX2 | Neuronal pentraxin-2 |
| NPTXR | Neuronal pentraxin receptor |
| NPY | Neuropeptide Y |
| NRGN | Neurogranin |
| Oligo-SNCA | Alpha-synuclein (oligomeric) |
| PARK7 | Parkinsonism-associated deglycase / DJ-1 |
| PDGFRB | Platelet-derived growth factor receptor beta |
| PDLIM5 | PDZ and LIM domain protein 5 |
| PGF | Placental growth factor |
| PGK1 | Phosphoglycerate kinase 1 |
| POSTN | Periostin |
| PRDX6 | Peroxiredoxin-6 |
| PSEN1 | Presenilin-1 |
| pSNCA-129 | Phosphorylated alpha-synuclein (Ser129) |
| pTau-181 | Microtubule-associated protein tau, phosphorylated at Thr181 |
| pTau-217 | Microtubule-associated protein tau, phosphorylated at Thr217 |
| pTau-231 | Microtubule-associated protein tau, phosphorylated at Thr231 |
| pTDP43-409 | TAR DNA-binding protein 43, phosphorylated at Ser409 |
| PTN | Pleiotrophin |
| REST | RE1-silencing transcription factor |
| RUVBL2 | RuvB-like AAA ATPase 2 |
| S100A12 | S100 calcium-binding protein A12 |
| S100B | S100 calcium-binding protein B |
| SAA1 | Serum amyloid A-1 protein |
| SFRP1 | Secreted frizzled-related protein 1 |
| SFTPD | Surfactant protein D / Pulmonary surfactant-associated protein D |
| SLIT2 | Slit homolog 2 protein |
| SMOC1 | SPARC-related modular calcium-binding 1 |
| SNAP25 | Synaptosomal-associated protein 25 |
| SNCA | Alpha-synuclein (monomer) |
| SNCB | Beta-synuclein |
| SOD1 | Superoxide dismutase [Cu-Zn] |
| SQSTM1 | Sequestosome-1 |
| TAFA5 | Chemokine-like protein TAFA-5 |
| TARDBP | TAR DNA-binding protein 43 (TDP-43) |
| TEK | Tie-2 / Angiopoietin-1 receptor |
| TIMP3 | Metalloproteinase inhibitor 3 / Tissue inhibitor of metalloproteinase 3 |
| TNF | Tumor necrosis factor |
| TREM1 | Triggering receptor expressed on myeloid cells 1 |
| TREM2 | Triggering receptor expressed on myeloid cells 2 |
| UCHL1 | Ubiquitin C-terminal hydrolase L1 |
| VCAM1 | Vascular cell adhesion molecule 1 |
| VEGFA | Vascular endothelial growth factor A |
| VEGFD | Vascular endothelial growth factor D |
| VGF | VGF nerve growth factor inducible (neurosecretory protein) |
| VSNL1 | Vasodilator-stimulated phosphoprotein-like protein 1 / Visinin-like protein 1 |
| YWHAZ | 14-3-3 protein zeta/delta |

**Supplementary Table 2 – Autosomal dominant Alzheimer disease genetic variants represented in this cohort**

| **Gene** | **Variants** |
| --- | --- |
| Amyloid precursor protein (*APP*) | V717I, V717G, V717L, T719N, V717F, V715A, I716V |
| Presenilin-1 (*PSEN1*) | A79V, Y115C, Y115H, T116I, E120K, N135S, M139V, V142I, I143F, M146I, M146V, L153V, H163R, I168del (TATdel), I168dup, S170P, E184D, I202F, G206A, G206V, H214Y, A246E, L262F, P264L, P267S, R269H, R278I, F283L, E280G, P436S, Intron 4 |
